# Lipoprotein(a) metabolomic and proteomic features and atherosclerotic cardiovascular disease in young, healthy adults

**DOI:** 10.1101/2024.05.12.24307240

**Authors:** Sascha N. Goonewardena, Tomasz Jurga, Donald Lloyd-Jones, Dilna Damodaran, Bharat Thyagarajan, David R. Jacobs, Supriya Shore, Eric J. Brandt, Clary Clish, Kahraman Tanriverdi, Jane E. Freedman, Brian T. Emmer, John T. Wilkins, Ron Do, Vera A. Bittner, Robert S. Rosenson, Ravi V. Shah, Venkatesh L. Murthy

**Affiliations:** Division of Cardiovascular Medicine, Department of Internal Medicine, University of Michigan, Ann Arbor, Michigan, USA; Division of Cardiovascular Medicine, VA Ann Arbor Health System, Ann Arbor, Michigan, USA; Department of Pharmacy, VA Ann Arbor Health System, Ann Arbor, Michigan, USA; Department of Preventive Medicine, Northwestern University Feinberg School of Medicine, Chicago, IL, USA; Department of Laboratory Medicine and Pathology, University of Minnesota, Minneapolis, Minnesota, USA; University of Minnesota School of Public Health, Division of Epidemiology and Community Health, Minneapolis, MN, USA; Institute for Healthcare Policy and Innovation, University of Michigan, Ann Arbor, Michigan, USA; Broad Institute of MIT and Harvard, Cambridge, Massachusetts, USA; Vanderbilt Translational and Clinical Research Center, Cardiovascular Division, Vanderbilt University Medical Center, Nashville, TN, USA; Department of Internal Medicine, Division of Hospital Medicine, University of Michigan Medical School, Ann Arbor, Michigan, USA; Division of Cardiology, Department of Medicine Northwestern, University Feinberg School of Medicine, Chicago, Illinois, USA; The Charles Bronfman Institute for Personalized Medicine, Icahn School of Medicine at Mount Sinai, New York, NY, USA; Department of Medicine, Division of Cardiovascular Disease, University of Alabama at Birmingham, Birmingham, AL, USA; Metabolism and Lipids Program, Zena and Michael A. Wiener Cardiovascular Institute, Mount Sinai Heart, Icahn School of Medicine at Mount Sinai, New York, New York, USA

**Keywords:** Inflammation, lipoproteins, thrombosis, atherosclerosis, ASCVD, lipoprotein(a)

## Abstract

**Background:** Elevated lipoprotein (a) [Lp(a)] is associated with a higher risk of atherosclerotic cardiovascular disease (ASCVD) events. Although Lp(a) is a genetically determined risk factor, the metabolomic and proteomic features that may mediate or be associated with this risk are unknown.

**Methods:** In young, healthy Coronary Artery Risk Development in Young Adults (CARDIA) participants, we defined the relationships between year 7 (Y7) Lp(a) and metabolomic (n=563)/proteomic (n=184) features, derived quantitative Lp(a)-omic scores from these features, and related these quantitative scores to ASCVD phenotypes.

**Results:** CARDIA participants had a mean age of 32 years at Y7 in this study and a median follow-up of 27.1 years. In the overall cohort (n=3920), Y7 Lp(a) levels were associated prospectively with ASCVD phenotypes at year 25 (Y25), including hs-CRP, coronary artery calcification (CAC), and incident CHD. In the subcohort (n=2290) that had measurements of Lp(a), proteomics, and metabolomics, Y7 Lp(a) levels were associated with distinct proteomic and metabolomic features indicative of immune responses, lipoprotein metabolism, atherogenesis, and arginine/steroid biosynthesis. Using machine learning approaches, Lp(a) metabolomic, proteomic, and transomic quantitative scores were derived. The Y7 Lp(a) transomic score was more strongly associated with Y25 incident CAC (standardized β = 0.29, p=0.04), hs-CRP (standardized β = 0.18, p =0.0008), and incident any CHD (standardized β = 0.51, p = 0.05), than the Y7 Lp(a) concentration itself (no significant associations).

**Conclusions:** To our knowledge, this is the first study to identify relationships between Lp(a) and associated metabolomic/proteomic features in young, healthy adults and joint associations with ASCVD phenotypes. The multi-omics approaches employed here provide insight into the pathobiology of Lp(a)-driven ASCVD and enable more nuanced mechanistic risk assessment compared with Lp(a) concentrations alone.

## Introduction

Despite major therapeutic advances, atherosclerotic cardiovascular disease (ASCVD) remains the leading cause of morbidity and mortality worldwide.^1^ This is due to a continued high prevalence of traditional risk factors, including a lack of heart-healthy dietary practices, and an aging population.^2^ However, even after accounting for traditional risk factors, significant risk remains. Part of this risk is mediated by elevated levels of lipoprotein(a) [Lp(a)]. Lp(a) is an apolipoprotein B particle that is covalently attached to an apolipoprotein(a) glycoprotein. Elevated concentrations of Lp(a) increase the risk for ASCVD events in persons with and without coronary heart disease (CHD).^3–5^ Observational cohort studies, post hoc analyses of clinical trials, and Mendelian randomization studies suggest an independent, likely causal relationship between elevated Lp(a) levels and ASCVD risk.^6–9^ Although there are currently no approved pharmacological therapies that directly target elevated lipoprotein(a), several therapies are in development that reduce apolipoprotein(a) production by up to 80%–99%.^10^

Mechanistically, Lp(a) promotes ASCVD through multiple pathways, including atherogenesis, thrombosis, and inflammation.^11^ Intriguingly, studies suggest that on a per particle basis, Lp(a) is associated with more atherosclerosis than LDL implying that Lp(a) possesses additional atherogenic properties compared with LDL.^12^ Given that a major structural component of Lp(a) is encoded by a gene (*LPA*) that was duplicated from the plasminogen gene, it has been hypothesized that modulation of fibrinolysis could mediate the effects of Lp(a) on thrombosis.^13–15^ However, several studies in murine models and human cohorts have failed to demonstrate a clear effect of Lp(a) on fibrinolysis or venous thromboembolic disease.^16^ Several basic and observational studies have demonstrated a relationship between Lp(a) and inflammation.^17^ In asymptomatic middle-aged participants in MESA (Multi-Ethnic Study of Atherosclerosis), Lp(a) ASCVD risk and all-cause mortality were only observed in combination with an elevated hs-CRP (≥2 mg/L).^18^ More recently, a risk factor and context-dependent relationship between Lp(a) and inflammation has emerged.^19^ A study by Arnold and colleagues found that high levels of Lp(a) were associated with an increased risk of CHD events, regardless of the level of hs-CRP in individuals without CHD.^20^ However, in individuals with established CHD, Lp(a) was only associated with recurrent CHD events in those with high hs-CRP levels. Functional evidence linking Lp(a) to inflammation has been revealed in experimental models and clinical studies demonstrating that Lp(a) is the primary lipoprotein carrier of phosphocholine-containing oxidized phospholipids (OxPL), a damage-associated molecular pattern (DAMP) that can be recognized by pattern recognition receptors (PRRs) on immune and non-immune cells.^21,22^ In total, Lp(a) appears to influence multiple pathways in a context- and subject-specific manner to drive ASCVD.

Although it is appreciated that Lp(a) can work through distinct mechanisms to drive ASCVD, how these mechanisms directly link to heterogeneous ASCVD endpoints has remained elusive. To identify candidate pathways that might mediate the increased risk of ASCVD, we leveraged Lp(a) associated metabolomic and proteomic signatures in conjunction with the longitudinal follow-up of young, healthy adults in CARDIA and identified pathway-specific associations linking Lp(a) with ASCVD phenotypes. Specifically, we employed 563 metabolomic and 184 proteomic features (approximately 1.7 million measurements) in 2299 participants with Lp(a) measurements in the CARDIA study. Using machine learning approaches, we developed Lp(a) quantitative metabolomic and proteomic signatures and related these to ASCVD phenotypes and clinical outcomes. Overall, these studies are the first to derive Lp(a) metabolomic and proteomic quantitative scores and link them to ASCVD phenotypes in a young, healthy adult population, providing quantitative risk assessments and novel mechanistic insights through which Lp(a) could accelerate ASCVD.

## Materials and Methods

### Study participants and Lp(a) measurements

The CARDIA study is a prospective, observational cohort study of 5115 participants (white and Black, age 18-30 years, recruited in 1985 and 1986 from Birmingham, AL; Chicago, IL; Minneapolis, MN; Oakland, CA). At inception, there were 5115 participants enrolled at ages 18-30 years, balanced on age, education status, and self-reported sex and race. CARDIA participants have undergone in-person examinations at baseline (Year 0: Y0) and at Y2, Y5, Y7, Y10, Y15, Y20, Y25, Y30, and Y35. Retention rates among surviving participants at each in-person examination were 91%, 86%, 81%, 79%, 74%, 72%, 72%, 71%, and 67% (during the COVID-19 pandemic), respectively. 1156 individuals were excluded due to missing Y7 Lp(a), 36 were excluded for missing key covariates (age, sex, race, total cholesterol, HDL-C LDL-C, triglycerides, diabetes status, systolic and diastolic blood pressure, use of antihypertensive medications, and smoking status at Y7), and 2 for incident coronary heart disease before Y7 resulting in an analysis sample of n=3920 (full cohort with Lp(a) measured at Y7) and n=2299 (subcohort with Lp(a) and metabolomics/proteomics at Y7) Participants provided written informed consent, and CARDIA was approved by Institutional Review Boards at participating institutions. Plasma Lp(a) analyses were performed by an ELISA method insensitive to apo[a] size heterogeneity.^23^

### Metabolite and proteomic profiling

We used plasma samples for metabolite and proteomic profiling. Metabolite profiling in CARDIA from Y7 samples has been previously described.^24^ In brief, the methods include a C8 reversed-phase liquid chromatography (LC) with positive ion mass spectrometry (MS) for lipid quantification (C8-pos); a C18 reversed-phase LC with negative ion MS (C18-neg) for quantification of fatty and bile acids; a hydrophilic interaction LC with positive ion MS (HILIC-pos) and negative ion MS (HILIC-neg) for cationic and anionic polar metabolites, respectively. For analyses, we included metabolites with missingness of <10% and CV<10%. Metabolite values were natural logarithm transformed to improve normality. We performed targeted proteomics assays (Olink CVD II and CVD III panels, Uppsala, Sweden) on Y7 samples in CARDIA.^25^ This proximity-extension assay technique generates amplification cycle numbers that are normalized (Olink NPX Manager Software v3.1.1.399) to protein expression levels (NPX) (arbitrary units on log2 scale).

### CARDIA Outcomes definitions

Contact is maintained with participants via telephone, mail, or email every 6 months, with annual interim medical history ascertainment. Over the last 5 years, >90% of the surviving cohort members have been directly contacted and follow up for vital status is virtually complete through related contacts and intermittent National Death Index searches. For this study, we evaluated clinical events (incident hard CHD, any CHD, and death from any cause as adjudicated per CARDIA protocol), and subclinical coronary atherosclerosis as measured by Agatston CAC scores on computed tomography at year 15, 20 and 25.^26^ CAC scores were treated either continuously (after transformation as the natural log transformation of the score plus 1) or dichotomized (as zero or non-zero). Hs-CRP was measured at Y7, year 15, year 20, and year 25. Hard CHD was defined as non-fatal myocardial infarction, non-fatal non-MI acute coronary syndrome, definite fatal MI, and definite or possible fatal CHD. For, “any CHD” coronary revascularization is added to the hard CHD composite of outcome. Time to events was defined from the Y7 examination date with a median follow-up of 27.1 years.

### Statistical analysis

We compared continuous and categorical variables across groups using Wilcoxon rank-sum and chi-square tests, respectively. We graphically evaluated distributions and elected to natural log transform Lp(a) to improve normality. Additionally, we converted Lp(a) to a binary variable by dichotomization at 150 nmol/L and used quintiles for further analyses. The high Lp(a) concentration was defined as ≥150 nmol/L (70 mg/dL) based on the inclusion criteria of the Lp(a)HORIZON clinical trial. We used linear, logistic, Cox, and Poisson regression for continuous, binary, time to event, and event rate outcomes, respectively. We scaled key continuous predictors and outcomes to unit variance and zero mean to present standardized beta coefficients. These regressions were performed without adjustment (unadjusted), adjusted for age/sex/race, or fully adjusted (for age, sex, race, total cholesterol, HDL-C LDL-C, triglycerides, diabetes status, systolic and diastolic blood pressure, use of antihypertensive medications, and smoking status [all at Y7]). Event rates for incident hard and any CHD and all-cause mortality across quintiles of Lp(a), LDL-C, and hs-CRP were computed with Poisson regression across each of the three adjustment approaches. Significance was evaluated for individual quintile groups as well as for linear trends across quintiles. We also related Lp(a) continuously, dichotomized at 150 nmol/L or across quintiles with all clinical and subclinical endpoints using linear, logistic, or Cox regression, as appropriate.

For metabolomic and proteomic analyses, we randomly divided the subset of the cohort with measured proteomics and metabolomics into 70% for derivation and 30% for validation samples. We performed linear regression with natural log transformed and standardized Lp(a) as the predictor and each proteomic and metabolomic feature as an outcome variable (also standardized). These were performed as unadjusted, age/sex/race adjusted, and fully adjusted models to derive standardized betas. P-values were adjusted using the Benjamini-Hochberg false discovery rate (FDR) control method within each predictor set and adjustment approach. These relationships were used to identify potentially mechanistically relevant relationships between Lp(a) and candidate effectors.

We also developed signatures of Lp(a) (continuous variable) using Least Absolute Shrinkage and Selection Operator (LASSO) penalized regression using metabolites features alone, proteomics features alone, or both combined as predictors. The LASSO hyperparameter (λ) was optimized with repeated 5-fold cross-validation. The signatures were fit in the (70%) derivation subset and then computed in both the derivation and validation subsets. Omics signatures were then related to the outcomes using linear, logistic, and Cox regression as above. Models including omics scores and additionally adjusted for Lp(a) were also fit to enable comparison of the scores with measured Lp(a). Statistical analyses were performed in R version 4.3.2 (R Foundation for Statistical Computing, Vienna, Austria), and a type 1 error of 5% (significance level 0.05) was used with type 1 error handling where noted.

Unadjusted metabolites and proteins with regression coefficients significant at a 5% FDR were selected for pathway analysis using MetaboAnalyst 6.0 (https://www.metaboanalyst.ca/) and mapped to both metabolic pathways and gene-only pathways in KEGG database, respectively. A hypergeometric test was used for enrichment analysis and degree centrality was used for topology measure. P-values from metabolites and proteins were combined via Z-test as an integrative enrichment P-value for each pathway. All pathways with an FDR less than 0.001 (in the pathway analysis for cross-sectional or longitudinal data) were selected for result interpretation and visualization.

### Resources, data, and code availability

Data have been deposited at the CARDIA Coordinating Center as listed in the key resources table. Access can be obtained directly from the CARDIA Coordinating Center (www.uab.dopm.edu/cardia) and is available as of the date of publication. Accession identifier numbers are listed in the key resources table. This paper does not report the original code. The code here uses standard packages in R studio. Any additional information required to reanalyze the data reported in this paper is available from the lead contact upon request.

## Results

### Characteristics of the CARDIA cohort

Lp(a) and key covariates were measured at year 7 (Y7) in 3920 participants without prior CHD. The demographic and laboratory characteristics of this cohort are shown in **Table 1**. The distribution of age and sex were similar across Lp(a) strata (**Table 1**). As previously described, Lp(a) levels were modestly higher in black compared with white participants.^27^ Lp(a) itself was only weakly correlated with other Y7 parameters including total cholesterol (R=0.14; p<0.0001), LDL-C (R=0.16; p<0.0001), HDL-C (R=0.05; p=0.004), triglycerides (R=-0.04; p=0.01), and hs-CRP (R=0.11; p<0.0001) (**Supplemental Figure 1**).

**Table 1.** Demographic and laboratory characteristics of CARDIA Lp(a) sample at Year 7, overall and stratified by Lp(a) level. The present study analyzes 3920 individuals with Lp(a) levels measured at Year 7 and complete assessment of key covariates (age, sex, race, diabetes, lipids, blood pressure, BMI, and smoking) and without CHD before year 7.

| Characteristic | N | Overall,<br>N=3,920 | Plasma Lp(a) <<br>150 nmol/L, N<br>= 3,377 | Plasma Lp(a) ≥<br>150 nmol/L, N =<br>543 | p-value |
| --- | --- | --- | --- | --- | --- |
| Age (y) | 3,920 | 32.0 (29.0, 35.0) | 32.0 (29.0, 35.0) | 32.0 (29.0, 35.0) | 0.9 |
| Male Sex | 3,920 | 1,767 (45%) | 1,528 (45%) | 239 (44%) | 0.6 |
| Race | 3,920 |  |  |  | <0.001 |
| Black |  | 1,884 (48%) | 1,513 (45%) | 371 (68%) |  |
| White |  | 2,036 (52%) | 1,864 (55%) | 172 (32%) |  |
| BMI (kg/m<sup>2</sup>) | 3,844 | 25.4 (22.6, 29.5) | 25.3 (22.5, 29.2) | 25.9 (23.0, 30.3) | 0.008 |
| Total Plasma Cholesterol (mg/dl) | 3,920 | 174 (153, 198) | 172 (152, 195) | 185 (166, 208) | <0.001 |
| HDL Cholesterol (mg/dl) | 3,920 | 50 (42, 60) | 50 (42, 60) | 51 (44, 61) | 0.028 |
| LDL Cholesterol (mg/dl) | 3,920 | 105 (85, 127) | 102 (84, 125) | 117 (97, 137) | <0.001 |
| Triglycerides (mg/dl) | 3,920 | 67 (47, 101) | 67 (47, 102) | 66 (47, 97) | 0.4 |
| Diabetes | 3,920 | 129 (3.3%) | 108 (3.2%) | 21 (3.9%) | 0.4 |
| Systolic BP (mmHg) | 3,920 | 107 (100, 115) | 107 (100, 115) | 108 (102, 116) | 0.048 |
| Diastolic BP (mmHg) | 3,920 | 69 (63, 75) | 68 (63, 75) | 69 (62, 76) | 0.5 |
| BP Medication Use | 3,920 | 72 (1.8%) | 58 (1.7%) | 14 (2.6%) | 0.2 |
| Smoking | 3,920 |  |  |  | 0.017 |
| never |  | 2,264 (58%) | 1,958 (58%) | 306 (56%) |  |
| former |  | 621 (16%) | 551 (16%) | 70 (13%) |  |
| current |  | 1,035 (26%) | 868 (26%) | 167 (31%) |  |
| Glucose (mg/dl) | 3,900 | 90 (85, 96) | 89 (85, 95) | 90 (86, 96) | 0.046 |
| Y7 CRP (ug/ml) | 3,831 | 1.1 (0.5, 3.2) | 1.1 (0.5, 3.1) | 1.4 (0.6, 3.5) | 0.002 |
| Y15 CRP (ug/ml) | 3,222 | 1.4 (0.6, 3.7) | 1.3 (0.6, 3.5) | 1.9 (0.7, 4.8) | <0.001 |
| Y20 hs-CRP (ug/ml) | 3,068 | 1.12 (0.48, 3.05) | 1.09 (0.47, 2.90) | 1.37 (0.58, 3.83) | <0.001 |
| Y25 hs-CRP (ug/ml) | 3,027 | 1.4 (0.6, 3.4) | 1.3 (0.6, 3.3) | 1.7 (0.7, 4.4) | <0.001 |
| Plasma Lp(a) (nmol/L) | 3,920 | 45 (14, 104) | 33 (12, 76) | 196 (167, 233) | <0.001 |
| Y15 CAC>0 | 2,732 | 299 (11%) | 254 (11%) | 45 (13%) | 0.2 |
| Y20 CAC>0 | 2,766 | 564 (17%) | 477 (17%) | 87 (21%) | 0.081 |
| Y25 CAC>0 | 2,782 | 904 (27%) | 752 (25%) | 152 (33%) | 0.003 |
| Incident Any CHD | 3,920 | 145 (3.7%) | 118 (3.5%) | 27 (5.0%) | 0.09 |
| Incident Hard CHD | 3,920 | 118 (3.0%) | 96 (2.8%) | 22 (4.1%) | 0.13 |
| Death | 3,920 | 313 (8.0%) | 268 (7.9%) | 45 (8.3%) | 0.8 |
| <sup>1</sup> Median (IQR); n (%) |  |  |  |  |  |
| <sup>2</sup> Wilcoxon rank sum test;<br>Pearson's Chi-squared<br>test |  |  |  |  |  |

### Lp(a) is associated with clinical markers of inflammation, atherosclerosis, and CHD events in young, healthy adults

To understand the relationship between Lp(a) concentrations and the risk of subclinical and clinical ASCVD, we studied participants in CARDIA with a median follow-up of 27.1 years. We quantified the risk of ASCVD (and associated atherogenic and inflammatory phenotypes) across the spectrum of observed Lp(a) concentrations as well as through Lp(a) quintiles. With respect to atherogenesis and inflammation, Y7 Lp(a) was associated with CAC and hs-CRP (**Figure 1** and **Supplemental Figure 2**). In the model adjusted for age, sex, and race, Lp(a) as a continuous variable was associated with incident hard and any CHD (**Figure 1**). The incidence of any and hard CHD was 1.82 per 1000 person-years and 1.43 per 1000 person-years in the highest quintile of Lp(a); in contrast, the incidence of any and hard CHD was 1.03 per 1000 person-years and 0.93 per 1000 person-years in the lowest quintile of Lp(a) (**Supplemental Figure 3**). Interestingly, the Lp(a) quintile analysis showed an inverse association between all-cause mortality in the age, sex, and race-adjusted model (**Supplemental Figure 2**). This association with mortality was not apparent when Lp(a) was examined using 150 nmol/L cutoff or Lp(a) as a continuous variable, nor was it significant in the unadjusted or fully adjusted models. To contextualize risk gradient across quintiles of Lp(a), we also conducted similar quintile analyses for Y7 LDL cholesterol (LDL-C) and hs-CRP and how these relate to CHD outcomes. With respect to any and hard CHD events, for the highest quintile of LDL-C, the incidence was 3.13 per 1000 person-years and 2.37 per 1000 person-years, respectively (**Supplemental Figure 3**), as compared to the lowest quintile of LDL-C. For hs-CRP, those in the highest quintile had an incidence of any and hard CHD events of 1.75 per 1000 person-years and 1.5 per 1000 person-years, respectively, more closely approximating the risk of the highest quintile of Lp(a) than LDL-C. In young, healthy adults, Lp(a) levels are associated not only with CHD endpoints, but also with atherogenic and inflammatory phenotypes.

**Figure 1.**
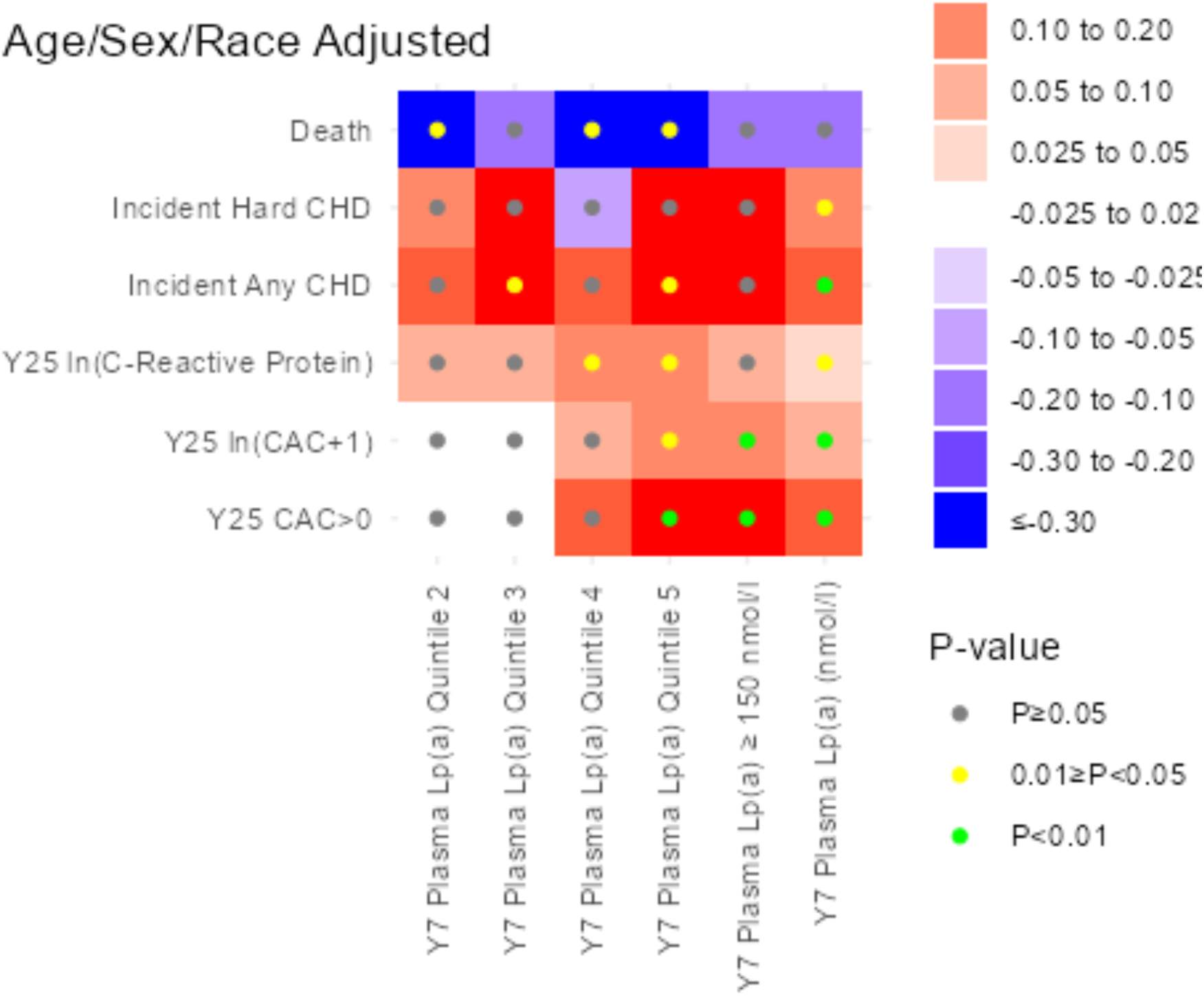
Y7 Lp(a) levels in young, healthy adults are associated with inflammatory and atherogenic phenotypes as well as CHD. Heatmap of standardized regression coefficients from models relating Y7 Lp(a) variables (quintiles, 150 nmol/L threshold, and as a continuous variable) to CAC, hs-CRP, and CHD endpoints (outcome variable). Models are adjusted for age, sex, and race. Values for regression coefficient directionality (color), magnitude (color gradient), and statistical significance (color of circle) are depicted in figure legend.

### Lp(a) levels in young, healthy adults are associated with distinct metabolic and proteomic features

Although Lp(a) influences ASCVD through multiple mechanisms, significant heterogeneity exists through which these pathological mechanisms contribute to the distinct clinical manifestations of ASCVD. To further illuminate the biological pathways that could underlie the associations between Lp(a) and ASCVD, we measured circulating metabolites and proteins in young adulthood (Y7 CARDIA) and how Y7 Lp(a) levels relate to these molecular features. The characteristics of this CARDIA subcohort (having Y7 Lp(a), metabolomics, and proteomics; N=2299) are shown in **Supplemental Table 1**. 563 metabolomic, and 184 proteomic features were analyzed in this subcohort. The results of linear, logistic, and Cox models are presented in the **Supplementary Data File** and **Figure 2**. From proteins and metabolites that were significant in single regression analyses (Y7 Lp(a) as the predictor variable), we found pathway annotations for 222 unique metabolites and 57 proteins. **Figure 2** shows a summary of the standardized regression coefficients for regression analyses across prioritized pathways. Pathway analyses identified cytokine-cytokine receptor interaction, lipid and atherosclerosis, arginine and proline metabolism, and steroid biosynthesis pathways which were related to Y7 Lp(a) levels.

**Figure 2.**
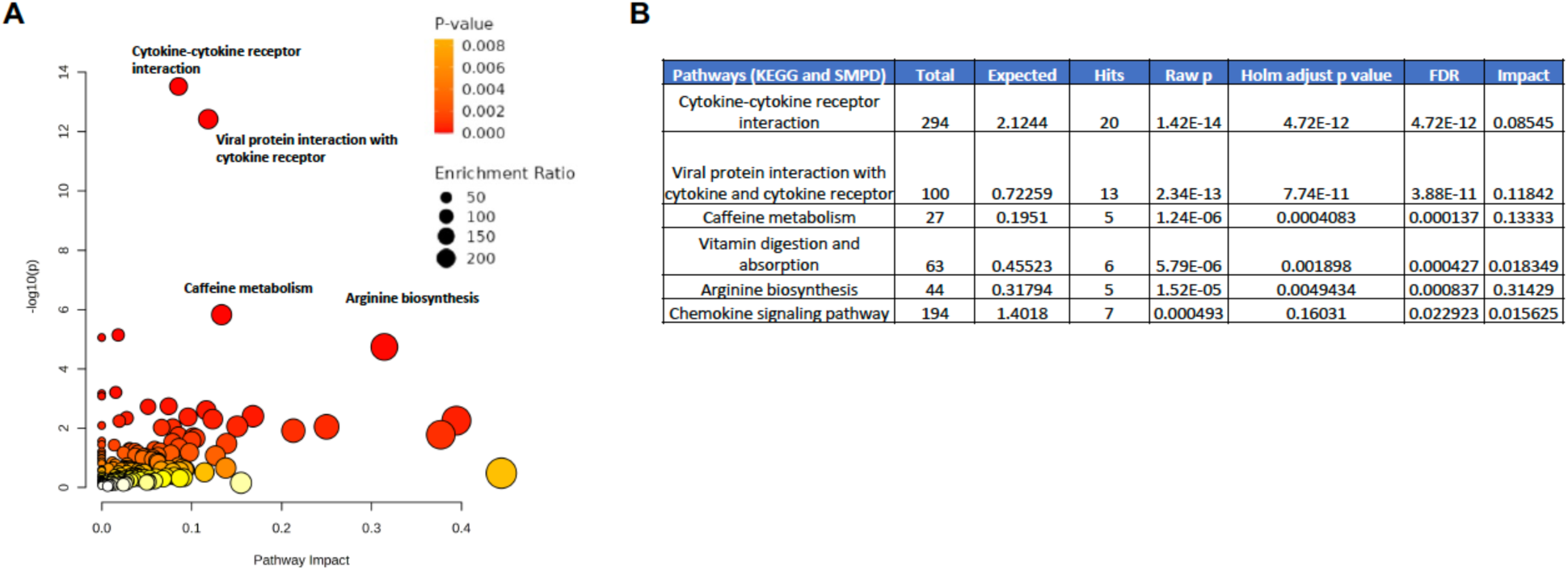
Lp(a) as a predictor of metabolomic and proteomic features. 222 unique metabolites and 57 proteomic features were selected using single regression based on FDR p-value <0.05. A) The dot plot of pathway topology result, where the X-axis shows the pathway impact score, and Y-axis are –log10(p) values. The size of the dot reflects the enrichment ratio, and the color reflects the level of significance. B) Top 6 significant pathways from both 85 KEGG mapped metabolites and 57 significant protein features.

### Derivation of Lp(a)-associated metabolomic, proteomic, and transomic signatures in young, healthy adults

Using LASSO regression analysis, we derived Lp(a)-quantitative scores comprised of the most significant metabolites and proteins (integration of them both [transomic]) (**Figure 3**). Not surprisingly, in the validation cohort, the metabolomic, proteomic, and transomic scores were significantly correlated with Y7 Lp(a) levels, from which they were derived, with correlation coefficients of 0.30, 0.25, and 0.30, respectively (**Supplemental Figure 4**). When examining the specific components of the metabolomic and proteomic scores that drive the association with Y7 Lp(a), several observations are worth noting. With respect to specific metabolites, some of the strongest contributions (both positive and negative) to the Lp(a) metabolic score were C24:1 ceramide, cytosine, and adenosine (**Figure 3** and **Supplemental Figure 5**). For specific proteins, some of the strongest contributions (both positive and negative) to the Lp(a) proteomic score included TFPI, MMP2/12, SCF, LPL, and NT-proBNP, which have all been linked to ASCVD (**Figure 3** and **Supplemental Figure 5**). These findings suggest that metabolic derangements, vascular dysfunction, and immune activation contribute to the Lp(a)-associated molecular instruments, providing insight into the pathobiology through which Lp(a) could underlie and accelerate ASCVD.

**Figure 3.**
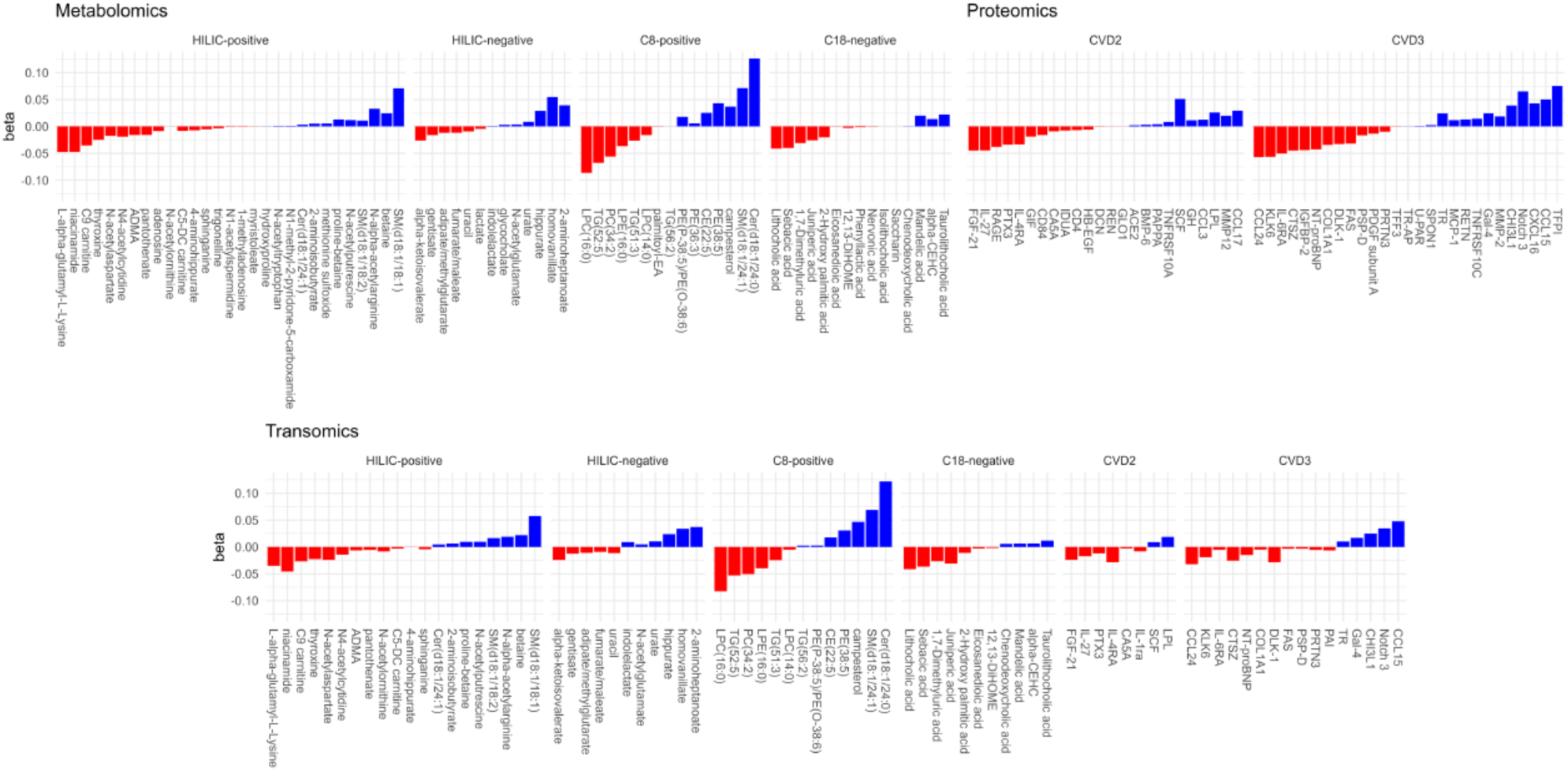
LASSO derived Lp(a) metabolomic, proteomic, and transomic scores and individual metabolomic/proteomic feature contributions. Standardized regression coefficients for metabolites and proteins are depicted as bars with the length of the bar proportionate to the weight, red color for negative weights, and blue for positive weights. Bars are grouped based on the metabolomic or proteomic platform used.

### Lp(a) derived metabolic and proteomic signatures are more strongly associated with accelerated atherogenesis and inflammation than Lp(a) itself

Because of the complex relationships between Lp(a), pathogenic pathways, and ASCVD clinical and subclinical phenotypes, we leveraged the quantitative Lp(a) omic scores. We assessed how these quantitative scores relate to clinical markers of inflammation (hs-CRP), atherosclerosis (CAC), and CHD outcomes. Although the Lp(a) omic scores were significantly correlated with Lp(a) levels, the correlation was modest, allowing for the possibility that the scores themselves may provide additional insights into Lp(a)-mediated ASCVD and ASCVD risk. In brief, we employed a statistical learning approach in which atherogenic (CAC), inflammatory (hs-CRP), and ASCVD events were modeled as a function of the Y7 Lp(a) metabolomic, proteomic, and transomic quantitative scores (**Figure 4** and **Supplemental Figure 6**). Because of the smaller number of participants in our CARDIA-omics subcohort (n=2299), not surprisingly, the significance of the associations between Lp(a) and clinical and subclinical ASCVD endpoints were muted compared with the larger cohort (n=3920). In this young, healthy subcohort, measured Lp(a) itself had minimal associations with ASCVD endpoints compared with Lp(a)-omics scores. For example, when comparing the Y7 Lp(a) transomic score to Y7 Lp(a) in the adjusted model, Y7 Lp(a) had minimal significant associations with ASCVD phenotypes. Specifically, the Y7 Lp(a) transomic score was more strongly associated with Y25 incident CAC (standardized β = 0.29, p=0.04), hs-CRP (standardized β = 0.18, p =0.0008), and incident any CHD (standardized β = 0.51, p = 0.05), than Y7 Lp(a) itself (no significant associations) (**Figure 4** and **Supplemental data**). Even with a reduced sample size in the subcohort, each of the Lp(a) -omic scores were significantly associated with ASCVD phenotypes in contrast with Lp(a) levels themselves, thereby providing provocative insight not only into the pathobiology of Lp(a) mediated ASCVD but also to ASCVD risk.

**Figure 4.**
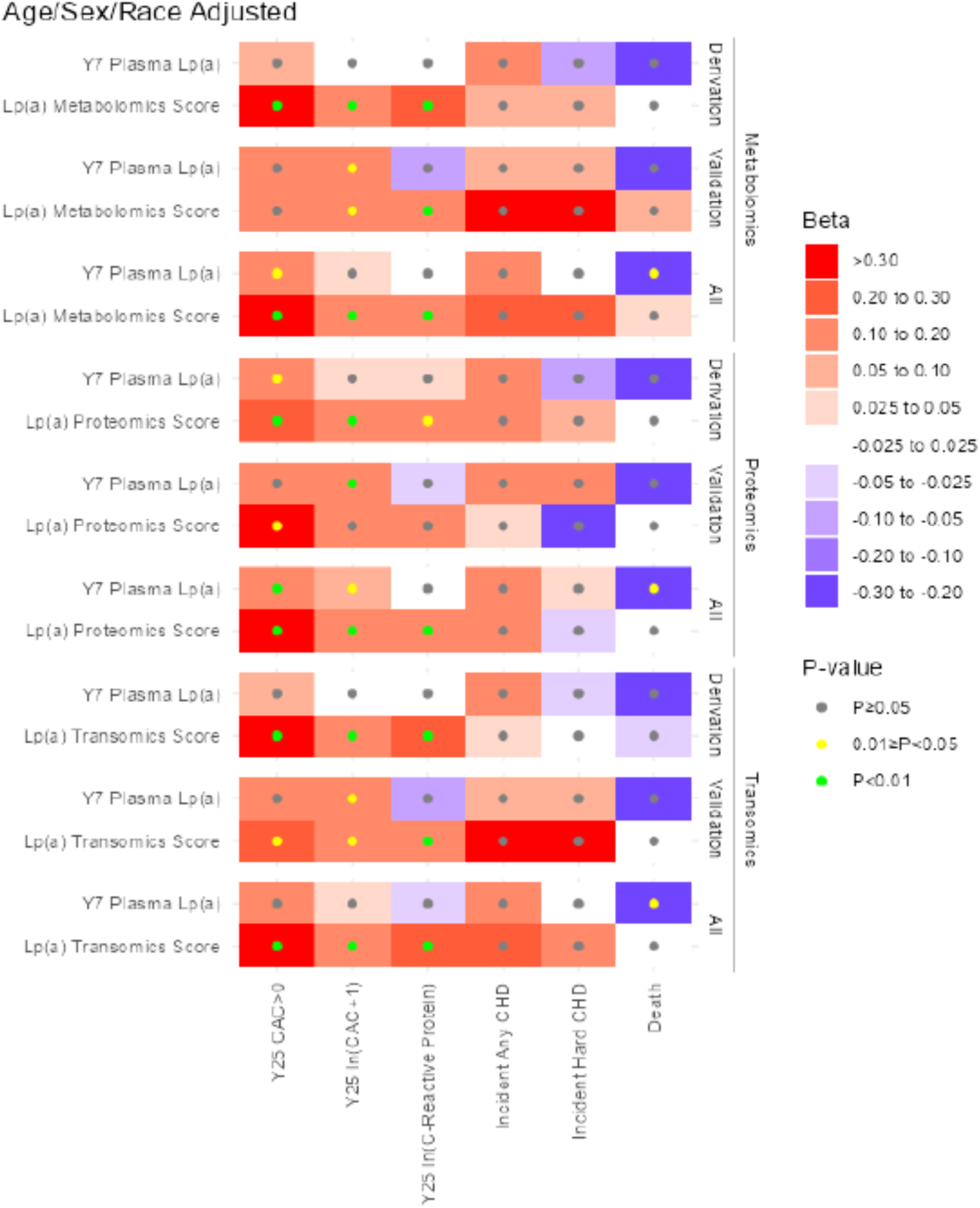
Lp(a) omics scores compared with measured Lp(a) as predictors of clinical and subclinical ASCVD endpoints. Heatmap depicting standardized beta relating omics scores for Lp(a) and measured Lp(a) to coronary artery calcification, high-sensitivity CRP, and outcomes using linear, logistic, and Cox regression adjusted for age, sex, and race. Results are grouped for the metabolomics score, proteomics score, and transomic score. Within these groups, regressions were separately performed in the score derivation set and score validation set and across both sets pooled (labeled All). Regressions were performed with measured Lp(a), and an omics score in the same model as predictors. Values for regression coefficient directionality (color), magnitude (color gradient), and statistical significance (color of circle) are depicted in figure legend.

## Discussion

In this study of young, healthy adults in CARDIA, we examined the relationships between Lp(a) levels and metabolomic/proteomic features and how these related to the development of subclinical and clinical ASCVD phenotypes. Our major findings are that: 1) Lp(a) levels in a young, healthy cohort are associated with accelerated atherogenesis, inflammation, and premature CHD endpoints; 2) Lp(a) levels are associated with specific metabolomic and proteomic features; 3) quantitative Lp(a) derived metabolomic and proteomic scores are associated with accelerated inflammation (hs-CRP), atherogenesis (CAC), and ASCVD phenotypes; and 4) the quantitative Lp(a) derived metabolomic and proteomic scores are more strongly related to these phenotypes than Lp(a) itself (from which these quantitative scores were derived). These findings are the first to link Lp(a) to metabolomic/proteomic features and to use the features to understand Lp(a) associated subclinical and clinical ASCVD phenotypes, providing potential mechanistic insight into the pathways through which Lp(a) accelerates ASCVD.

Extending on prior findings from higher-risk populations, we found that in young, healthy individuals, Lp(a) is associated with subclinical ASCVD phenotypes and CHD risk. Specifically, Lp(a) was associated not just with incident CHD but also with atherogenic phenotypes (e.g., year 25 CAC) and evidence of heightened inflammation (e.g., hs-CRP at year 25). A study of middle-aged participants in the UK Biobank also demonstrated that higher Lp(a) concentrations were associated with an increased risk of ASCVD events.^27^ This differential risk (between participants with a high and low Lp(a)) was more pronounced in individuals without a prior history of ASCVD. Additionally, a study of participants from the UK Biobank, as well as additional analyses of participants from the FOURIER and SAVOR-TIMI 53 trials, showed that higher Lp(a) is associated with a higher risk of ASCVD in both primary and secondary prevention populations regardless of baseline hs-CRP.^28^ One unique aspect of our study is the evaluation of a younger patient population with less cardiovascular risks, namely less hypertension, diabetes, and lower baseline LDL-C when compared to MESA, ARIC, and the UK Biobank.^18,29–31^ In higher-risk populations, LDL-C and hs-CRP have been associated with ASCVD phenotypes. Consistent with these findings, we also found that LDL-C and hs-CRP are associated with CHD and Lp(a) risk is more similar to hs-CRP with respect to the magnitude of ASCVD risk. These findings carry major implications given that Lp(a) is less dynamic than other markers of ASCVD risk. For example, markers such as LDL-C respond to lifestyle modifications and drug therapy, while Lp(a) remains largely unchanged.^32^ Intriguingly, perhaps because of the lower ASCVD risk of the CARDIA cohort, there was a trend towards increased mortality in the lowest quintile of Lp(a), which is discordant from ASCVD events. The total number of deaths overall is low, and the magnitude and significance were not dependent on Lp(a) levels. Future studies are needed to verify this unexplained finding which could be due to competing risk or simply due to chance. Overall, our results extend the ASCVD Lp(a) findings from other observation cohorts to a younger, lower-risk population, connecting Lp(a) to markers of inflammation and atherogenesis and ASCVD phenotypes.

Clinical trials with RNA-based therapeutics that reduce Lp(a) levels by 80-99% are ongoing.^10^ They will directly test the “Lp(a) hypothesis” that profound reductions in Lp(a) levels will reduce the risk of ASCVD. To complement these clinical outcomes trials, understanding the underlying mechanisms through which Lp(a) accelerates ASCVD and how individual-specific risk influences these mechanisms will not only help in identifying patients most likely to benefit from these therapeutics but also in expanding our knowledge of ASCVD itself. Our metabolomic and proteomic findings reinforce known and reveal unknown relationships between Lp(a) and pathways that underlie ASCVD. Among the correlates of Lp(a) in our single regression analyses, we identified several markers of vascular dysfunction (RAGE, PSGL1) and inflammation (MERTK, MCP1), all of which have been associated with ASCVD.^33–36^ With respect to metabolites, the links to Lp(a) in this young, healthy cohort were strong. For example, C9 carnitine and SM(d18:1/24:1) were associated with Y7 Lp(a) levels, and both have been linked to ASCVD and ASCVD risk factors, suggesting that Lp(a) could be modulating metabolic risk upstream of these features.^37,38^ From a pathway perspective, the metabolomic and proteomic features revealed both known and unknown pathways through which Lp(a) could be driving ASCVD. The immune-mediated pathways (e.g. cytokine-cytokine receptor interaction, chemokine signaling) jointly implicated from the metabolomic and proteomic have clear links to ASCVD.^39^ Arginine biosynthesis is a provocative pathway associated with Lp(a) metabolomic features and has clear links to ASCVD.^40^ As alluded to earlier, the multi-omic approaches not only provide insight into quantitative, patient-specific risk but also provide insight into the mechanisms that give rise to ASCVD heterogeneity and the multitude of pathways that could accelerate ASCVD.

Using metabolic and proteomic features, we created quantitative Lp(a) metabolomic, proteomic, and transomic scores and related these quantitative scores to ASCVD phenotypes. ASCVD heterogeneity is a manifestation of the complex convergent processes that accelerate ASCVD. With respect to the pathobiology of ASCVD, Lp(a) has multifarious properties that increase atherogenesis, thrombosis, and inflammation, all of which can accelerate ASCVD and trigger atherothrombotic events. Evidence of this can be seen in our data, in that Lp(a) is associated with pathway-specific molecular features and clinical markers of inflammation (hs-CRP) and atherogenesis (CAC). The individual Lp(a) proteomic and metabolomic features themselves had minimal relationships to ASCVD phenotypes. However, the machine learning quantitative scores, a composite of the individual metabolomic/proteomic features, were not only associated with ASCVD phenotypes, but these associations were stronger than the level of Lp(a) itself.

## Study Limitations

Several limitations of our study warrant consideration. First, although our clinical assay for Lp(a) concentration employs an isoform-insensitive immunoassay, we did not have information on oxidized phospholipids content that may more precisely link metabolomic/proteomic features to distinct Lp(a) properties. Additionally, our design was a split-set design, as the ASCVD risk factor distribution (e.g., age and diabetes) and metabolite/protein coverage are unique to this cohort. Although the uniqueness of our cohort and data set captures an understudied early timepoint in Lp(a) driven ASCVD risk, these findings must be validated in other populations with a higher prevalence of ASCVD risk factors. Finally, the observational nature of our study precludes any strict determinations of causality with respect to Lp(a) pathobiology; however, we believe that our findings are hypothesis generating and can help inform future mechanistic studies.

## Conclusions

In summary, we integrated metabolomics and proteomics with Lp(a) levels in young, healthy adults to reveal molecular mediators through which Lp(a) may drive ASCVD. More broadly, the metabolomic and proteomic approaches employed here provide insight into ASCVD heterogeneity and how patient-specific exposures interact with Lp(a) during the genesis of ASCVD. This study adds further evidence linking inflammation and atherogenesis in ASCVD and furthers our understanding whereby Lp(a) can be used to identify individuals at risk for ASCVD.

## Data Availability

All data produced in the present study are available upon reasonable request to the authors.

## AUTHOR CONTRIBUTIONS

## Acknowledgments

Although not directly involved in the conduct of this study, we thank the volunteers, patients, and staff caring for them for their contribution and commitment to clinical research.

## Abbreviations

ASCVD: atherosclerotic cardiovascular disease
CAC: coronary artery calcification
CARDIA: Coronary Artery Risk Development in Young Adults
CVD: cardiovascular disease
CHD: coronary heart disease
DAMP: damage-associated molecular pattern
hs-CRP: high-sensitivity C-reactive protein
LDL: low-density lipoprotein
Lp(a): Lipoprotein(a)
MESA: Multi-Ethnic Study of Atherosclerosis
OxPL: oxidized phospholipids
PRRs: pattern recognition receptors
Y7: year 7

