## Supplemental materials for "Lipoprotein(a) metabolomic and proteomic features and atherosclerotic cardiovascular disease in young, healthy adults"

**Contents**

1. Supplemental data preparation and analyses.

2. Supplemental Table 1. CARDIA -omics subcohort demographics and laboratory values

3. Supplemental Figure 1. Lp(a) and correlations with lipid and inflammatory parameters.

4. Supplemental Figure 2. Lp(a) and ASCVD endpoints; full CARDIA cohort.

5. Supplemental Figure 3. LDL-C, hs-CRP, Lp(a) comparative quintile analysis.

6. Supplemental Figure 4. Correlation between Lp(a) derived -omic scores and Y7 plasma Lp(a).

7. Supplemental Figure 5. Lp(a) associated pathway analysis for proteomic and metabolomic features.

8. Supplemental Figure 6. Lp(a) quantitative scores and subclinical and clinical ASCVD outcomes.

**Supplemental data preparation and analyses.**

Pathway analysis of metabolomics was performed using	MetaboAnalyst	6.0. The pathways analysis on the 85 significant metabolites selected from model variable Y7	Plasma	Lp(a) (nmol/l)	from unadjusted models was performed in MetaboAnalystR	6.0. The pathways analysis based on the HMDB ID of significant metabolites was completed with the Pathway Analysis module based on the default quantitative enrichment analysis method and the human KEGG database. Over-representation analysis (ORA) is performed when a list of compound names is provided (HMDB ID compound names). The list of compounds can be obtained through a clustering algorithm or from the compounds with abnormal concentrations detected in single sample profiling (SSP) to investigate if some biologically meaningful patterns were identified. ORA was implemented using the hypergeometric test to evaluate whether a particular metabolite set is represented more than expected by chance within the given compound list. One-tailed p values are provided after adjusting for multiple testing.

Proteomics pathway analysis was performed using R (4.2.3) libraries clusterProfiler, KEGGREST, and org.Hs.eg.db for a comprehensive exploration of biological pathways associated with differentially expressed proteins. The raw proteomics data were preprocessed and normalized using methods. Followed by statistical testing and FDR. Subsequently, the differentially expressed proteins were selected based on their P value (adjusted p-value < 0.05). To understand the biological significance of these proteins, pathway enrichment analysis was performed utilizing the clusterProfiler package. This package facilitates the functional annotation of gene clusters by linking them to known biological pathways and processes. KEGG pathway information was retrieved using the KEGGREST library, and gene-to-pathway mappings were obtained from the org.Hs.eg.db database. The enriched pathways were then visualized using ggplot2. The significance threshold for pathway enrichment was set at a pre-defined significance level (p-value < 0.05).

**Supplemental Table 1.** **Demographic and laboratory characteristics of CARDIA Lp(a) multi-omics sample at Year 7, overall and stratified by Lp(a) level.** The omics study analyzes 2299 individuals with Lp(a) levels measured at Year 7 and complete assessment of key covariates (age, sex, race, diabetes, lipids, blood pressure, BMI, and smoking), metabolomics and proteomics measures and without CHD before Year 7.

| **Characteristic** | **N** | Overall, N = 2,299 | Derivation, N = 1,609 | Validation, N = 690 | p-value |
| --- | --- | --- | --- | --- | --- |
| Age (y) | 2,299 | 33.0 (29.0, 35.0) | 32.0 (29.0, 35.0) | 33.0 (30.0, 35.0) | 0.2 |
| Male Sex | 2,299 | 1,260 (55%) | 882 (55%) | 378 (55%) | >0.9 |
| Race | 2,299 |  |  |  | 0.8 |
| Black |  | 1,026 (45%) | 716 (44%) | 310 (45%) |  |
| White |  | 1,273 (55%) | 893 (56%) | 380 (55%) |  |
| BMI (kg/m2) | 2,276 | 24.8 (22.4, 27.9) | 24.8 (22.4, 28.0) | 24.8 (22.4, 27.7) | 0.6 |
| Total Plasma Cholesterol (mg/dl) | 2,299 | 174 (154, 196) | 174 (155, 195) | 173 (153, 196) | 0.9 |
| HDL Cholesterol (mg/dl) | 2,299 | 52 (44, 62) | 52 (44, 62) | 52 (44, 62) | 0.8 |
| LDL Cholesterol (mg/dl) | 2,299 | 103 (85, 126) | 103 (85, 126) | 103 (84, 126) | >0.9 |
| Triglycerides (mg/dl) | 2,299 | 63 (45, 91) | 63 (46, 92) | 64 (45, 90) | 0.4 |
| Diabetes | 2,299 | 48 (2.1%) | 42 (2.6%) | 6 (0.9%) | 0.007 |
| Systolic BP (mmHg) | 2,299 | 107 (101, 114) | 107 (101, 115) | 107 (100, 114) | 0.3 |
| Diastolic BP (mmHg) | 2,299 | 68 (63, 74) | 68 (63, 74) | 68 (62, 74) | 0.3 |
| BP Medication Use | 2,299 | 24 (1.0%) | 15 (0.9%) | 9 (1.3%) | 0.4 |
| Smoking | 2,299 |  |  |  | 0.6 |
| never |  | 1,402 (61%) | 975 (61%) | 427 (62%) |  |
| former |  | 367 (16%) | 265 (16%) | 102 (15%) |  |
| current |  | 530 (23%) | 369 (23%) | 161 (23%) |  |
| Glucose (mg/dl) | 2,292 | 90 (85, 95) | 90 (85, 95) | 89 (84, 94) | 0.5 |
| Y7 CRP (ug/ml) | 2,290 | 0.90 (0.42, 2.40) | 0.89 (0.42, 2.40) | 0.93 (0.41, 2.41) | 0.6 |
| Y15 CRP (ug/ml) | 1,980 | 1.2 (0.5, 3.2) | 1.2 (0.5, 3.0) | 1.3 (0.5, 3.3) | 0.3 |
| Y20 hsCRP (ug/ml) | 1,901 | 1.01 (0.45, 2.52) | 1.01 (0.45, 2.49) | 1.01 (0.46, 2.56) | 0.8 |
| Y25 hsCRP (ug/ml) | 1,866 | 1.2 (0.6, 2.9) | 1.3 (0.6, 2.9) | 1.2 (0.6, 2.9) | >0.9 |
| Plasma Lp(a) (nmol/l) | 2,299 | 39 (13, 99) | 38 (13, 99) | 43 (14, 95) | 0.7 |
| Plasma Lp(a) Quintile | 2,299 |  |  |  | 0.2 |
| Q1 |  | 509 (22%) | 362 (22%) | 147 (21%) |  |
| Q2 |  | 485 (21%) | 344 (21%) | 141 (20%) |  |
| Q3 |  | 437 (19%) | 304 (19%) | 133 (19%) |  |
| Q4 |  | 439 (19%) | 288 (18%) | 151 (22%) |  |
| Q5 |  | 429 (19%) | 311 (19%) | 118 (17%) |  |
| Plasma Lp(a) ≥ 150 nmol/l | 2,299 | 300 (13%) | 219 (14%) | 81 (12%) | 0.2 |
| Y15 CAC>0 | 1,696 | 170 (10%) | 120 (10%) | 50 (9.6%) | 0.7 |
| Y20 CAC>0 | 1,994 | 329 (16%) | 226 (16%) | 103 (17%) | 0.7 |
| Y25 CAC>0 | 2,105 | 543 (26%) | 378 (26%) | 165 (26%) | 0.7 |
| Incident Any CHD | 2,299 | 71 (3.1%) | 47 (2.9%) | 24 (3.5%) | 0.5 |
| Incident Hard CHD | 2,299 | 53 (2.3%) | 34 (2.1%) | 19 (2.8%) | 0.3 |
| Death | 2,299 | 141 (6.1%) | 90 (5.6%) | 51 (7.4%) | 0.1 |
| *^1^* Median (IQR); n (%) | | | | | |
| *^2^* Wilcoxon rank sum test; Pearson’s Chi-squared test | | | | | |

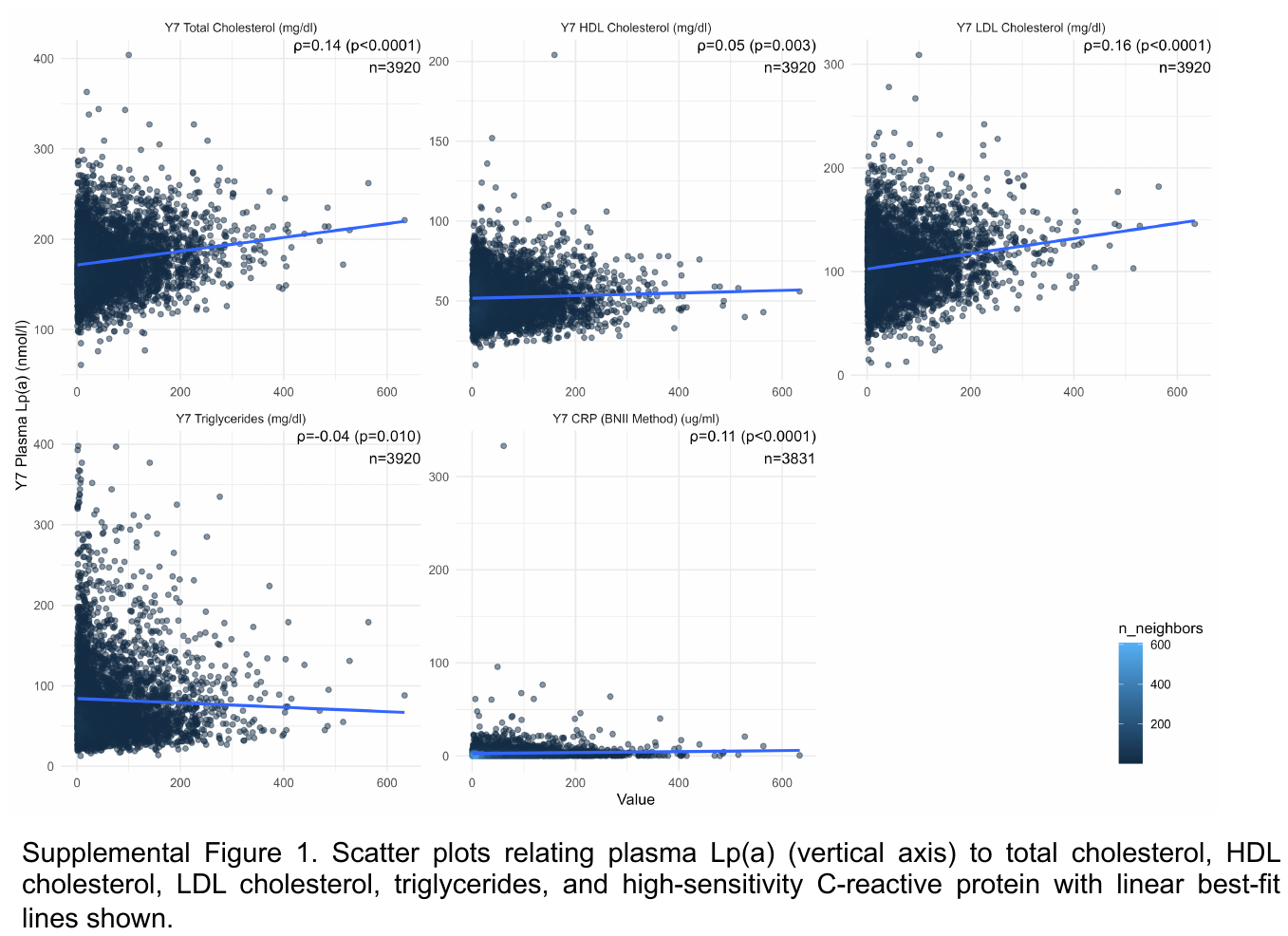

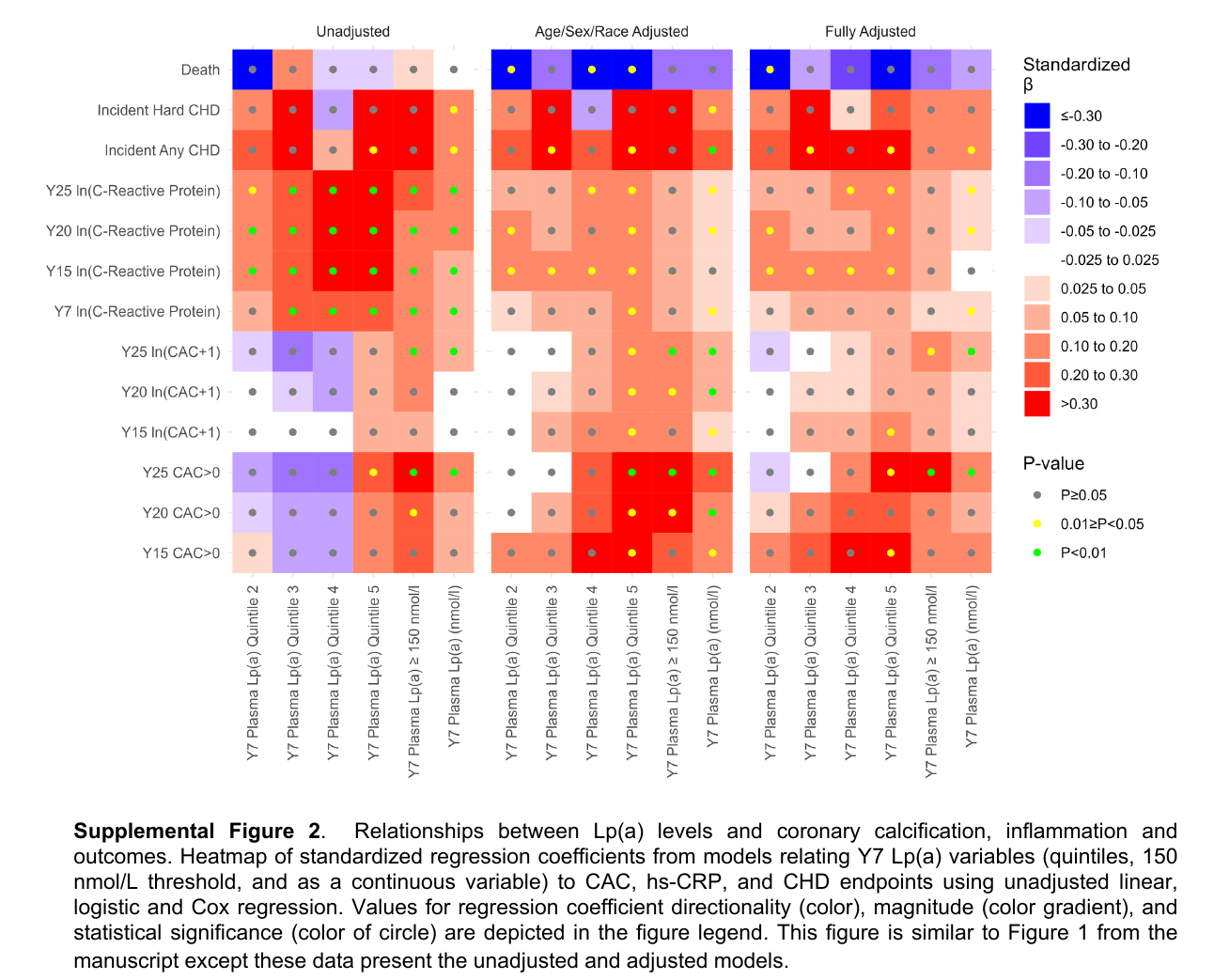

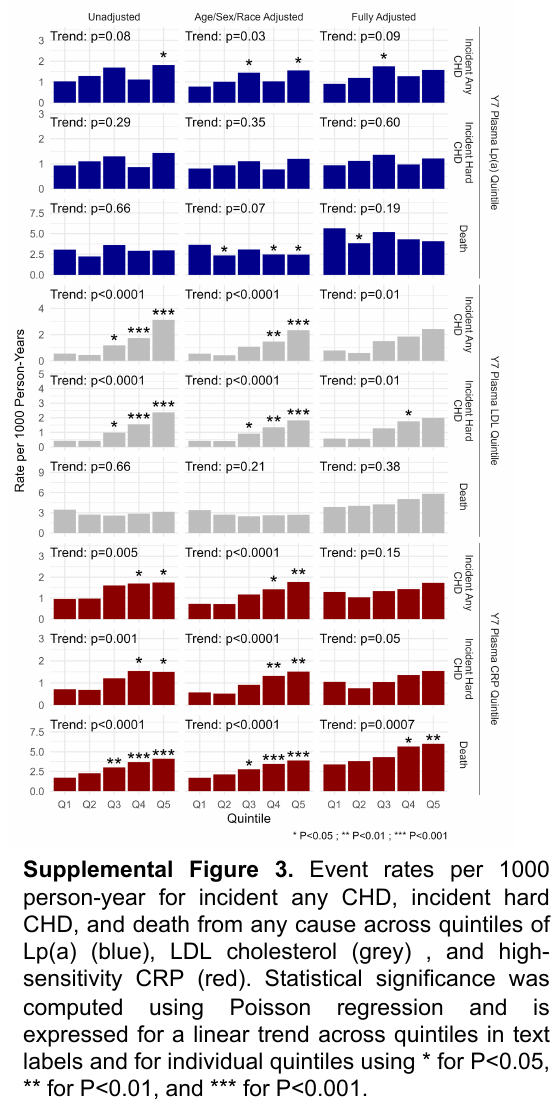

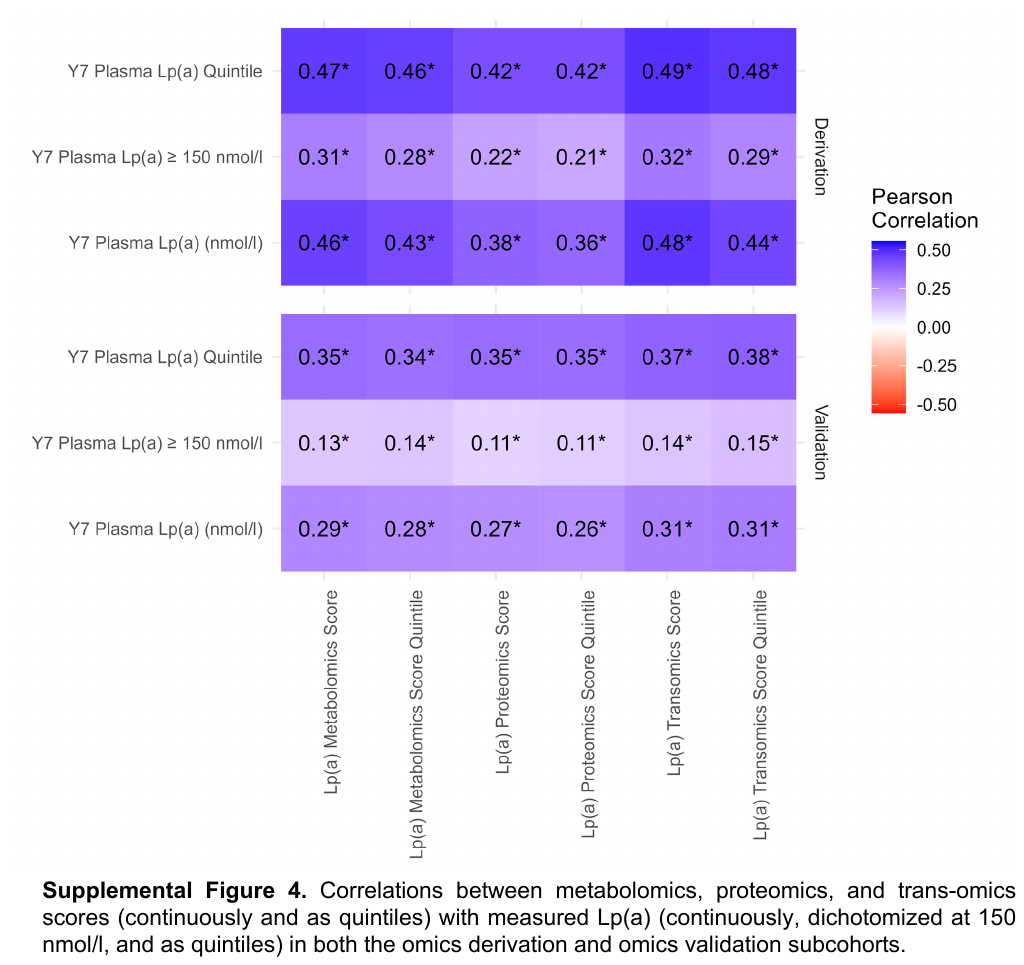

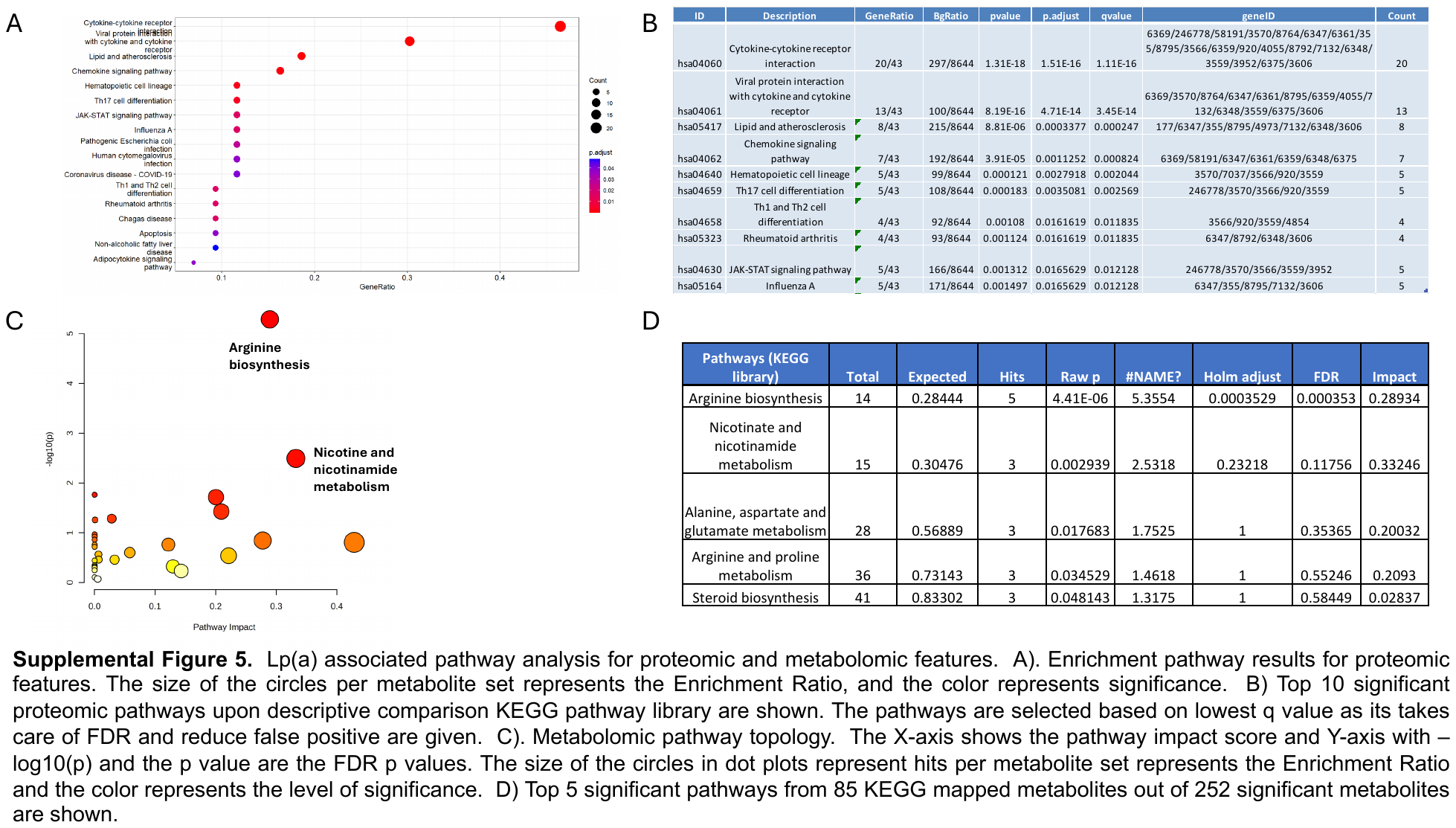

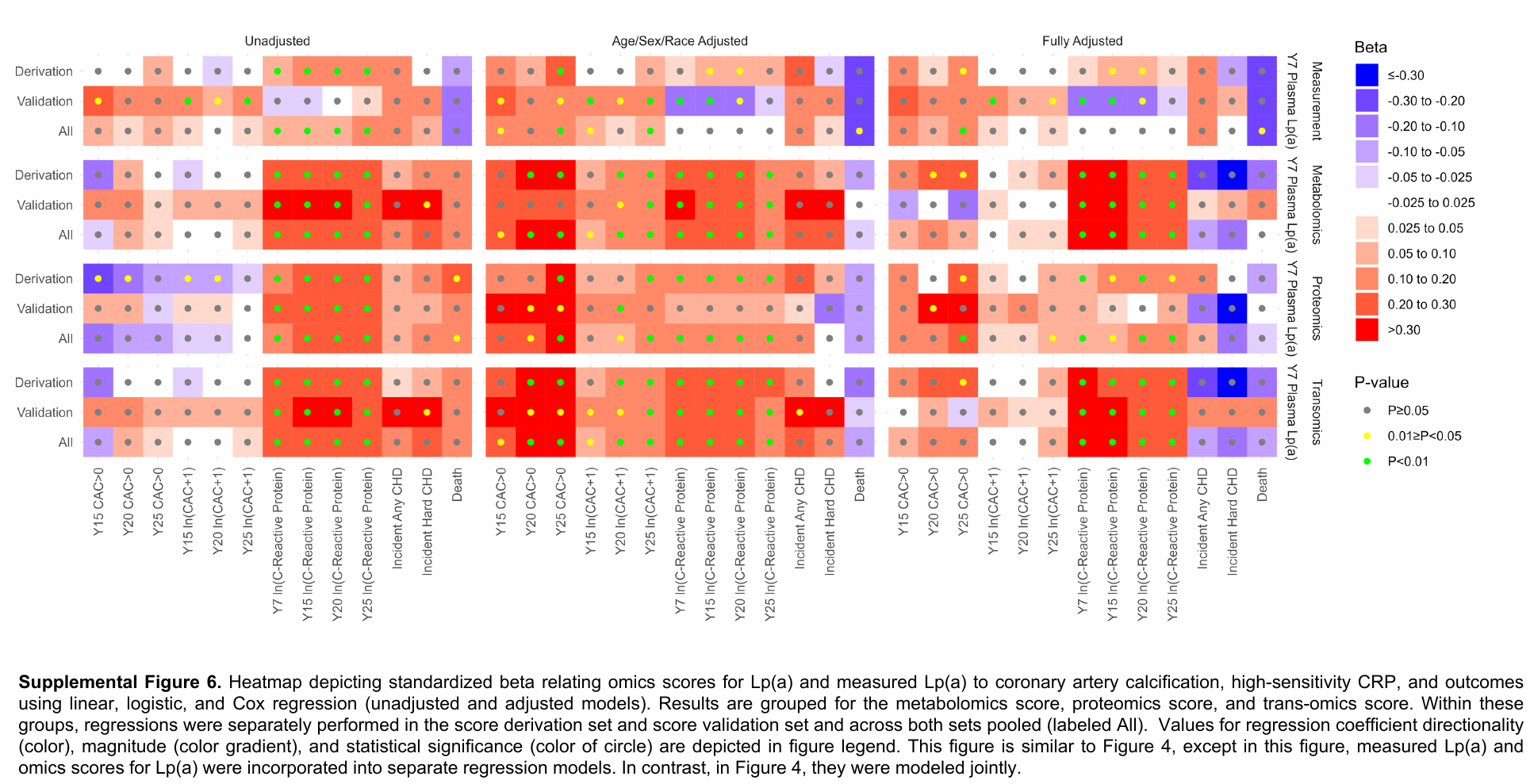
